# Psychomotor Vigilance Test and Epworth Sleepiness Scale in Participants being Evaluated for Sleep Disorders

**DOI:** 10.1101/2025.06.03.25328917

**Authors:** Allison Schwab, Brendan T. Keenan, Mathias Basner, Charles J. Bae

## Abstract

**Study Objectives:** Excessive daytime sleepiness (EDS) is common in participants with sleep disorders, particularly obstructive sleep apnea (OSA), and can be assessed using the Epworth Sleepiness Scale (ESS) and the Psychomotor Vigilance Test (PVT). However, the relationship between these measures of sleepiness/attention, and their relationships to OSA severity and treatment, remains understudied. This study examined these associations in a sleep center population.

**Methods:** A total of 167 participants, primarily diagnosed or suspected of OSA (n=128 [76.6%]), completed the ESS and PVT during their clinical visit. Associations among ESS, PVT, OSA severity and CPAP adherence were examined using Pearson’s correlations, unadjusted and controlling for age, sex and body mass index.

**Results:** Results showed no significant correlations between ESS and PVT measures of attention/vigilance. While higher ESS scores correlated with more severe apnea-hypopnea index (AHI) in participants with OSA, no association was found with PVT measures. Among participants using continuous positive airway pressure (CPAP), greater hours/night of usage was associated with lower ESS scores, but not with better PVT performance.

**Conclusions:** Our data indicate that ESS scores track more closely than PVT to OSA severity and treatment. The findings suggest that the tendency to fall asleep as measured by the ESS and attention deficits on PVT may capture different aspects of “sleepiness”. While the ESS is commonly used in sleep clinics, further research is needed to determine if PVT should also be used routinely in clinical practice.

**Brief summary:** Excessive daytime sleepiness (EDS) is a prevalent symptom among individuals with obstructive sleep apnea (OSA), but the relationship between a subjective measure (Epworth Sleepiness Scale) and an objective measure (Psychomotor Vigilance Test) of sleepiness or attention, as well as how each relates to OSA severity and treatment, is not well understood. This study found no association between the ESS and measures from a 3-minute PVT, suggesting that these assessments are not evaluating the same aspects of “sleepiness” reported by participants. Higher ESS scores, but not worse PVT performance, was related to more severe OSA and less adherence to CPAP, indicating that the ESS tracks more closely than the PVT to OSA severity and treatment use.

## Introduction

Among patients with sleep disorders, excessive daytime sleepiness (EDS) is a frequently reported symptom that can lead to a decrease in cognitive performance, mood, and overall wellbeing.^1^ EDS is commonly measured by the Epworth Sleepiness Scale (ESS), a self-reported questionnaire assessing the likelihood of dozing off or falling asleep across common activities^2,3^. While the ESS measures daytime sleepiness, it has also been used to screen participants for obstructive sleep apnea (OSA), as EDS is a very common symptom of OSA^4^. In participants with OSA, greater sleepiness on the ESS is associated with high blood pressure and cardiovascular disease risk^5–11^.

Although the ESS is the most commonly used metric for characterizing EDS in participants with OSA, it only has moderate utility to screen for OSA^12^. Data are also mixed on how well the ESS associates with OSA severity based on metrics such as the apnea hypopnea index (AHI)^13,14^. This suggests additional measures of EDS may help to more accurately quantify a patient’s sleepiness symptoms. For example, recent work has suggested that the addition of a single question on feeling sleepy during the day to the ESS can identify participants with worse quality of life and more OSA-related symptomatology^15^. There is also a growing literature on OSA symptom-based subtypes, one of which (termed *excessively sleepy*) is defined by the presence of multiple symptoms of excessive sleepiness and may account for the elevated cardiovascular risk related to OSA^6–8,10,16–19^. Notably, the ESS alone is insufficient to identify participants with this at-risk subtype^19,20^.

Measuring EDS objectively may provide a better representation of the impact of daytime sleepiness among participants with OSA. Towards this end, the psychomotor vigilance test (PVT) provides objective measures of deficits in attention and vigilance^21,22^. The PVT is a reaction time test that has been used to measure deficits of sleep loss, such as optimal neurobehavioral functioning in astronauts^23^, screening for impaired vigilance in commercial drivers and emergency responders^24^, and poor sleep quality affecting medical attentiveness in nurses^25,26^. The PVT is highly sensitive to small changes in attention, which can occur from sleep deprivation and other sleep disorders^27^. Additionally, the PVT has no learning curve and its reliability and validity have been proven by previous studies^28,29^.

Prior studies have found that the PVT and ESS are correlated, with higher ESS scores associated with a greater number of lapses in untreated participants with OSA^30–32^. However, these findings have not been replicated in a clinical sleep setting. To better understand the manifestation and measurement of “sleepiness”, this study examines the relationship between these two measures in a sample of participants from a clinical sleep center. We hypothesized that the ESS and PVT measures of sleepiness would be correlated, but the PVT (since it is an objective measure) would be more strongly correlated with AHI severity and adherence to continuous positive airway pressure (CPAP) than the subjective ESS. If this were the case, then sleep clinics should consider incorporating a PVT as a standard metric in participants being evaluated for OSA and treatment outcomes.

We will also examine correlations with the ESS and PVT to mask leak and residual AHI (based on PAP downloaded data) in the participants being treated with CPAP. We do not know if participants who have a large mask leak or a high residual AHI are sleepy. There are standard cutoffs for large leak (> 24 l/min on a ResMed device, > than 1 hour on a Philips device), and residual AHI (> 10 events/hour)^33^. It would be important to know if these clinical endpoints in mask leak or residual AHI are associated with sleepiness.

## Materials and Methods

### Subjects

This investigation is based on data gathered from a clinical sample of participants who were evaluated in the Sleep Center at the University of Pennsylvania over a 3-month period (6/2023 to 8/2023). The sample consisted of individuals being evaluated for sleep disorders who completed their PVT and ESS during their clinic visit. Additional information was extracted from the electronic health record (EHR). The exclusion criteria were missing information on both the PVT and ESS, due to either missing data (e.g., physician did not obtain information or patient did not have time to complete PVT at their visit) or an inability to complete the tasks (cognitive impairment, or language barrier). This project was reviewed and determined to qualify as Quality Improvement by the University of Pennsylvania’s Institutional Review Board.

### Epworth Sleepiness Scale (ESS)

Participants completed the ESS at the beginning of their clinical visit. The ESS includes eight questions asking the patient to rate their tendency to doze or fall asleep (on a scale of 0 to 3) across eight common and varied situations^2^. The total score of the ESS is calculated by adding up patient responses to each question and can range between 0 to 24, with a score > 10 commonly used to indicate excessive daytime sleepiness.

### Psychomotor Vigilance Test (PVT)

At the conclusion of the clinical visit, participants completed the 3-minute PVT^34^ on a HP Mobile Workstation ZBook 15 G3 15.6 inches FHD Laptop. The PVT is an attention/reaction time test in which participants are tasked with pressing a button as soon as a randomly-timed visual stimulus appears. The PVT measures the speed and accuracy with which participants respond to the stimuli, providing an objective measure of alertness. Primary measures from PVT include the number of lapses (defined as a response time [RT] >500ms), which is transformed as square root of lapses plus square root of lapses + 1 for analysis, and the mean reciprocal response time (mean RRT)^27^.

### Sleep Studies

Of the 167 participants, 142 had sleep studies, including 102 (71.8%) in-laboratory polysomnography (PSG; Philips G3) studies and 40 (28.2%) home sleep studies (HSAT). Twenty-five participants did not have sleep studies: 6 of those participants had insomnia and did not need a sleep study, 15 had a sleep study ordered but it was never completed, and 4 never had a sleep study ordered. During the PSG, apneas were defined as an absence of airflow for >10 seconds and hypopneas as a 50% reduction in airflow for >10 seconds associated with a ≥4% decrement in oxygen saturation and/or an arousal. During the HSAT (Philips NightOne or Clevemed) nasal airflow, chest and abdominal movements, pulse, oxygen saturation, body position and body movement were all measured.

### Continuous Positive Airway Pressure (CPAP)

Of the 128 participants with OSA, 47 (36.7%) were known to be on CPAP before their clinical visit; 26 were on ResMed devices, 20 on Philips devices, and 1 on a Resvent device. CPAP data were obtained on 42 (89.4%) of these 47 participants from Philips and ResMed databases, including the average usage (on all days and on days used) in the one month, one week, and one day prior to the visit, as well as the residual AHI (events per hour) and device-specific leak measures (time in large leak from Philips devices and 95^th^ percentile leak from ResMed devices) over the one month before the visit.

### Statistical Methods

Continuous variables are summarized using means and standard deviations (SDs) and categorical variables using frequencies and percentages. Where presented, comparisons of demographic variables between groups were performed using T-tests (continuous) or chi-squared and Fisher’s exact tests (categorical). To examine the associations between ESS and PVT measures, we utilized Pearson’s linear correlations and partial correlations adjusted for age, sex and body mass index (BMI). Associations were examined among all participants and separately in participants with and without OSA. Similar analyses were used to examine the associations between OSA severity and sleepiness measures among participants with OSA with available AHI not using CPAP, as well as to evaluate associations between CPAP usage, residual AHI, or leak metrics and sleepiness measures among participants with OSA in whom CPAP data were available. As defined by Cohen^35^, correlation coefficients of 0.1, 0.3 and 0.5 are interpreted as small, medium and large effect sizes. A Hochberg step-up procedure was utilized to determine statistical significances while maintaining type I error at 5% within the context of multiple comparisons performed in each of the above analyses^36,37^.

## Results

### Sample Characteristics

A total of 167 participants were included in this study (see **Table 1**). Participants were middle-aged (54.7±16.8 years), obese (BMI of 34.5±9.9 kg/m^2^), a majority were males (58.7%), and there were a similar proportion of Black (44.7%) and White (42.1%) participants. Of the 167 participants, 128 (76.6%) had diagnosed or suspected OSA. Participants with OSA were more likely to be male (p=0.001) and had higher BMI (p=0.0003) and older age (p=0.054) than those without OSA.

**Table 1.** Descriptive characteristics of the analysis sample.

| Measure | All | No OSA | OSA | p |
| --- | --- | --- | --- | --- |
| N | 167 (100%) | 39 (23.4%) | 128 (76.6%) | – |
| Age (years) | 54.7 ± 16.8 | 49.5 ± 19.5 | 56.3 ± 15.7 | 0.054 |
| Male | 58.7% | 35.9% | 65.6% | 0.001 |
| BMI (kg/m<sup>2</sup>) | 34.5 ± 9.9 | 30.3 ± 9.4 | 35.8 ± 9.7 | 0.003 |
| Weight (pounds) | 223.1 ± 66.3 | 188.4 ± 55.6 | 233.5 ± 65.8 | 0.0001 |
| Height (inches) | 67.5 ± 4.0 | 66.4 ± 3.7 | 67.8 ± 4.0 | 0.047 |
| Race |  |  |  |  |
| White | 42.1% | 58.3% | 37.4% | 0.123 |
| Black | 44.7% | 36.1% | 47.2% |  |
| Asian | 5.0% | 2.8% | 5.7% |  |
| Other | 8.2% | 2.8% | 9.8% |  |
| Sleep Diagnosis |  |  |  |  |
| OSA | 76.7% | 0.0% | 100.0% | n/a |
| Snoring | 7.2% | 30.8% | 0.0% |  |
| Insomnia | 8.4% | 35.9% | 0.0% |  |
| Idiopathic Hypersomnia | 1.8% | 7.7% | 0.0% |  |
| Parasomnias including RBD | 3.0% | 12.8% | 0.0% |  |
| Narcolepsy | 0.6% | 2.6% | 0.0% |  |
| Other* | 2.4% | 10.3% | 0.0% |  |
| AHI (events/hour) | 28.0 ± 28.1 | 1.58 ± 1.60 | 33.4 ± 27.9 | <0.0001 |
| SpO<sub>2</sub> nadir (%) | 80.6 ± 8.6 | 88.1 ± 5.8 | 79.0 ± 8.3 | <0.0001 |
| Minutes with SpO<sub>2</sub> < 90% | 30.9 ± 61.2 | 2.4 ± 6.8 | 37.0 ± 65.8 | <0.0001 |
| ESS | 8.07 ± 5.39 | 6.97 ± 5.00 | 8.39 ± 5.47 | 0.138 |
| PVT Lapses | 10.6 ± 10.7 | 10.9 ± 9.1 | 10.5 ± 11.2 | 0.828 |
| PVT Transformed Lapses | 5.97 ± 2.93 | 6.22 ± 2.61 | 5.89 ± 3.02 | 0.509 |
| PVT Mean RRT | 3.39 ± 0.69 | 3.36 ± 0.44 | 3.40 ± 0.75 | 0.682 |
| *Other Diagnoses (circadian rhythm disorders, poor sleep hygiene, myopathy); Abbreviations: BMI = Body Mass Index; AHI = Apnea Hypopnea Index; ODI = Oxygen Desaturation Index; OSA = Obstructive Sleep Apnea; RBD = REM behavior disorder; PVT = Psychomotor Vigilance Test; ESS = Epworth Sleepiness Scale; RRT = Reciprocal Response Time |  |  |  |  |

Similarly, characteristics of participants with diagnosed or suspected OSA stratified by whether or not the participant was using CPAP prior to their clinical visit are presented in **Table 2**. On average, the 47 (36.7%) participants using CPAP were about 10 years older (62.1 ±15.5 years) than those not using CPAP (52.9±14.9 years; p=0.002), but other demographics characteristics were generally similar. Those using CPAP had some evidence of less severe hypoxemia, but a higher average AHI (see **Table 2**). Out of the 47 participants using CPAP, objective adherence and efficacy data were available for 42 (89.4%) for analyses of the associations with sleepiness measures. These participants used CPAP for an average of 5.96±2.18 hours/night. The median (IQR) residual AHI was 3.0 (1.3, 5.2) events/hour, time in large leak for those on Philips devices was 13.9 (0.6, 93.6) minutes, and 95^th^ percentile leak for those on ResMed devices was 20.8 (8.6, 40.4) L/min.

**Table 2.** Descriptive characteristics of participants with OSA using and not using CPAP at the time of the clinical visit.

| Measure | OSA | No CPAP | Using CPAP* | p |
| --- | --- | --- | --- | --- |
| N | 128 (100%) | 81 (63.3%) | 47 (36.7%) | — |
| Age (years) | 56.3 ± 15.7 | 52.9 ± 14.9 | 62.1 ± 15.5 | 0.002 |
| Male | 65.6% | 64.2% | 68.1% | 0.655 |
| BMI (kg/m<sup>2</sup>) | 35.8 ± 9.7 | 36.2 ± 10.3 | 35.0 ± 8.8 | 0.485 |
| Weight (pounds) | 233.5 ± 65.8 | 237.1 ± 68.1 | 227.2 ± 61.9 | 0.410 |
| Height (inches) | 67.8 ± 4.0 | 68.0 ± 4.1 | 67.4 ± 3.8 | 0.437 |
| Race |  |  |  |  |
| White | 37.4% | 31.2% | 47.8% | 0.269 |
| Black | 47.2% | 53.3% | 37.0% |  |
| Asian | 5.7% | 5.2% | 6.5% |  |
| Other | 9.8% | 10.4% | 8.7% |  |
| AHI (events/hour) | 33.4 ± 27.9 | 30.1 ± 25.1 | 38.5 ± 31.3 | 0.132 |
| SpO<sub>2</sub> nadir (%) | 79.0 ± 8.3 | 77.7 ± 8.3 | 81.0 ± 7.9 | 0.038 |
| Minutes with SpO<sub>2</sub> < 90% | 37.0 ± 65.8 | 49.4 ± 77.7 | 16.8 ± 30.7 | 0.002 |
| ESS | 8.39 ± 5.47 | 9.00 ± 5.77 | 7.36 ± 4.82 | 0.089 |
| PVT Lapses | 10.5 ± 11.2 | 10.7 ± 11.9 | 10.0 ± 10.2 | 0.702 |
| PVT Transformed Lapses | 5.89 ± 3.02 | 5.96 ± 3.08 | 5.77 ± 2.96 | 0.725 |
| PVT Mean RRT | 3.40 ± 0.75 | 3.36 ± 0.81 | 3.47 ± 0.64 | 0.416 |
| *Data on average usage available in n = 42; Abbreviations: BMI = Body Mass Index; AHI = Apnea Hypopnea Index; ODI = Oxygen Desaturation Index; PVT = Psychomotor Vigilance Test; ESS = Epworth Sleepiness Scale; RRT = Reciprocal Response Time |  |  |  |  |

### Association between ESS and PVT

Participants had an average ESS of 8.1±5.4 and averaged 10.6±10.7 lapses (6.0±2.9 transformed lapses) and a mean RRT (reciprocal response time) of 3.4±0.7 seconds^−1^ on the PVT (see **Table 1**). Among all participants, there were no significant correlations between ESS and either transformed lapses (adjusted rho = 0.10, p=0.229) or mean RRT (adjusted rho = -0.09, p=0.277); results and interpretations were similar when comparing participants with or without diagnosed or suspected OSA (see **Table 3** and **Figure 1**).

**Figure 1.**
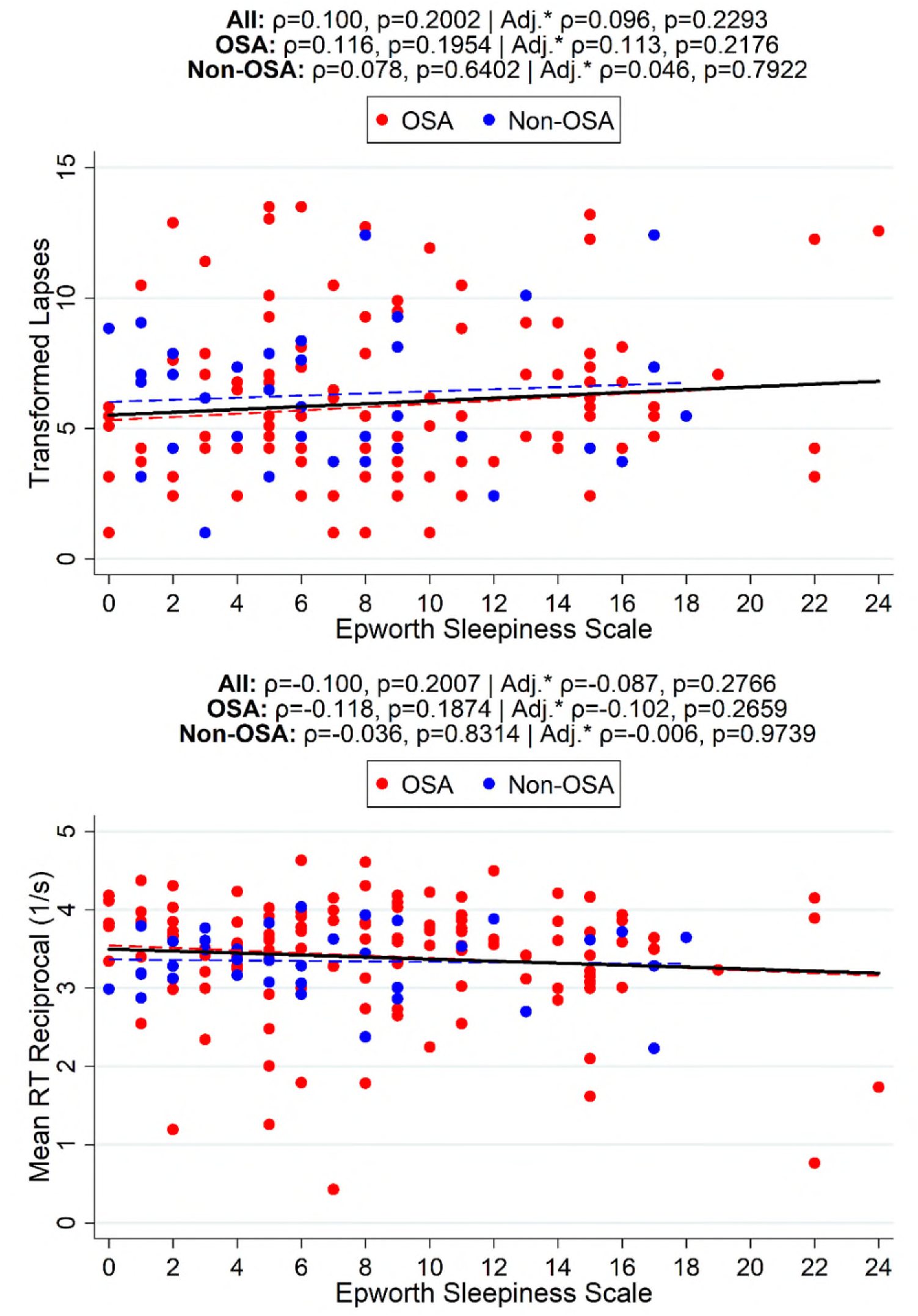
Associations among sleepiness measures and OSA severity. The associations between the Epworth Sleepiness Scale (ESS) and Psychomotor Vigilance Test (PVT) measures of (A) transformed lapses and (B) mean reciprocal response time (RRT) are illustrated in all participants and separately in those with and without OSA. The data showed no meaningful correlations between subjective sleepiness and either PVT measurement for OSA and Non-OSA participants (see also **Table 3**). *Adjusted for age, sex and BMI.

**Table 3.** Correlations between ESS and PVT measurements.

| PVT Measure | All Participants |  |  |  | Non-OSA |  |  |  | OSA |  |  |  |
| --- | --- | --- | --- | --- | --- | --- | --- | --- | --- | --- | --- | --- |
|  | *Unadjusted* |  | *Adjusted\** |  | *Unadjusted* |  | *Adjusted\** |  | *Unadjusted* |  | *Adjusted\** |  |
|  | *rho* | *p* | *rho* | *p* | *rho* | *p* | *rho* | *p* | *rho* | *p* | *rho* | *p* |
| Transformed Lapses | 0.10 | 0.200 | 0.10 | 0.229 | 0.08 | 0.640 | 0.05 | 0.792 | 0.12 | 0.195 | 0.11 | 0.218 |
| Mean RRT | -0.10 | 0.201 | -0.09 | 0.277 | -0.04 | 0.831 | -0.01 | 0.974 | -0.12 | 0.187 | -0.10 | 0.266 |
| *Partial correlation adjusted for age, sex and BMI; Abbreviations: PVT = Psychomotor Vigilance Test; ESS = Epworth Sleepiness Scale; RRT = Reciprocal Response Time |  |  |  |  |  |  |  |  |  |  |  |  |

### Associations with OSA Severity

Associations between measures of OSA severity and sleepiness were also evaluated among participants with OSA (see **Table 4** and **Figure 2**). More severe OSA was generally associated with higher ESS scores based on the AHI (adjusted rho = 0.28, p=0.025) or minutes SpO_2_ <90% (rho = 0.33, p=0.007); the association with AHI was not statistically significant after Hochberg correction but represents a moderate correlation. None of the OSA severity measures were significantly correlated with PVT-based measures of attention/vigilance.

**Figure 2.**
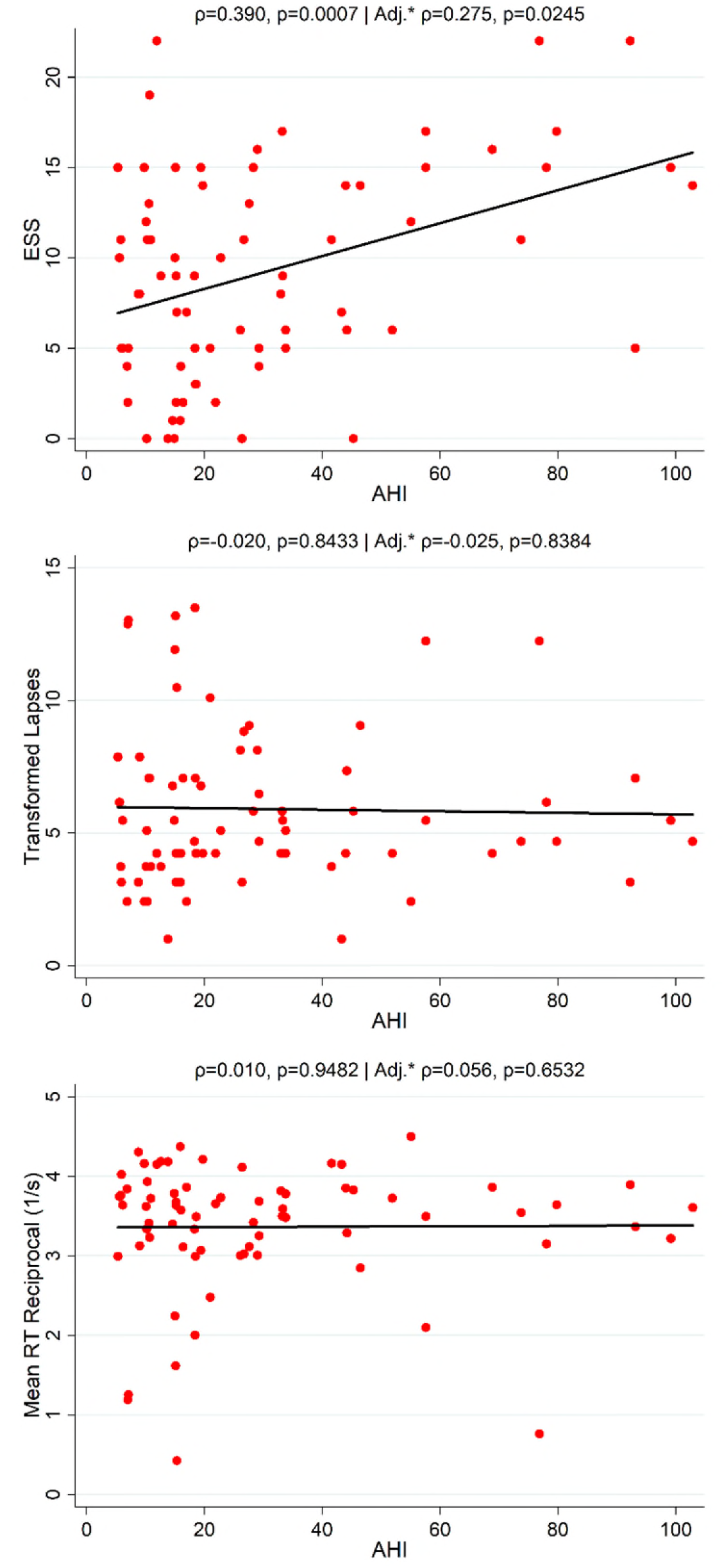
Associations between Apnea-Hypopnea Index (AHI) and sleepiness measures among participants with OSA not using CPAP. We observed significant correlations between AHI and subjective sleepiness based on ESS, but no relationship between AHI and objective measures of attention/vigilance from PVT (see also **Table 4**). *Adjusted for age, sex and BMI.

**Table 4.** Correlations between OSA Severity and both ESS and PVT measurements.

| Sleepiness Measure | AHI |  |  |  | SpO<sub>2</sub> nadir |  |  |  | Minutes SpO<sub>2</sub> < 90%<sup>†</sup> |  |  |  |
| --- | --- | --- | --- | --- | --- | --- | --- | --- | --- | --- | --- | --- |
|  | *Unadjusted* |  | *Adjusted\** |  | *Unadjusted* |  | *Adjusted\** |  | *Unadjusted* |  | *Adjusted\** |  |
|  | *rho* | *p* | *rho* | *p* | *rho* | *p* | *rho* | *p* | *rho* | *p* | *rho* | *p* |
| ESS | **0.39** | **0.001** | 0.28 | 0.025 | **-0.35** | **0.003** | -0.21 | 0.095 | **0.38** | **0.001** | **0.33** | **0.007** |
| Transformed Lapses | -0.02 | 0.843 | -0.03 | 0.838 | -0.19 | 0.115 | -0.19 | 0.130 | 0.15 | 0.220 | 0.13 | 0.307 |
| Mean RRT | 0.01 | 0.948 | 0.06 | 0.653 | 0.17 | 0.166 | 0.12 | 0.332 | -0.11 | 0.370 | -0.05 | 0.677 |
| Statistically significant associations after Hochberg correction for 3 sleepiness measures shown in **bold**. *Partial correlation adjusted for age, sex and BMI; Abbreviations: PVT = Psychomotor Vigilance Test; ESS = Epworth Sleepiness Scale; RRT = Reciprocal Response Time; <sup>†</sup>natural log transformed for analyses |  |  |  |  |  |  |  |  |  |  |  |  |

### Effects of CPAP Adherence on ESS Scores and PVT Performance

Among 42 participants with OSA with available data on amount of CPAP usage, greater hours/night of CPAP usage over the 30 days (adjusted rho = -0.47, p=0.003), 7 days (adjusted rho = -0.43, p=0.011), and the day prior to visit (adjusted rho = -0.32, p=0.050) were associated with lower ESS scores (see **Table 5** and **Figure 3**). No correlations were found between the amount of CPAP use and measures from PVT.

**Figure 3.**
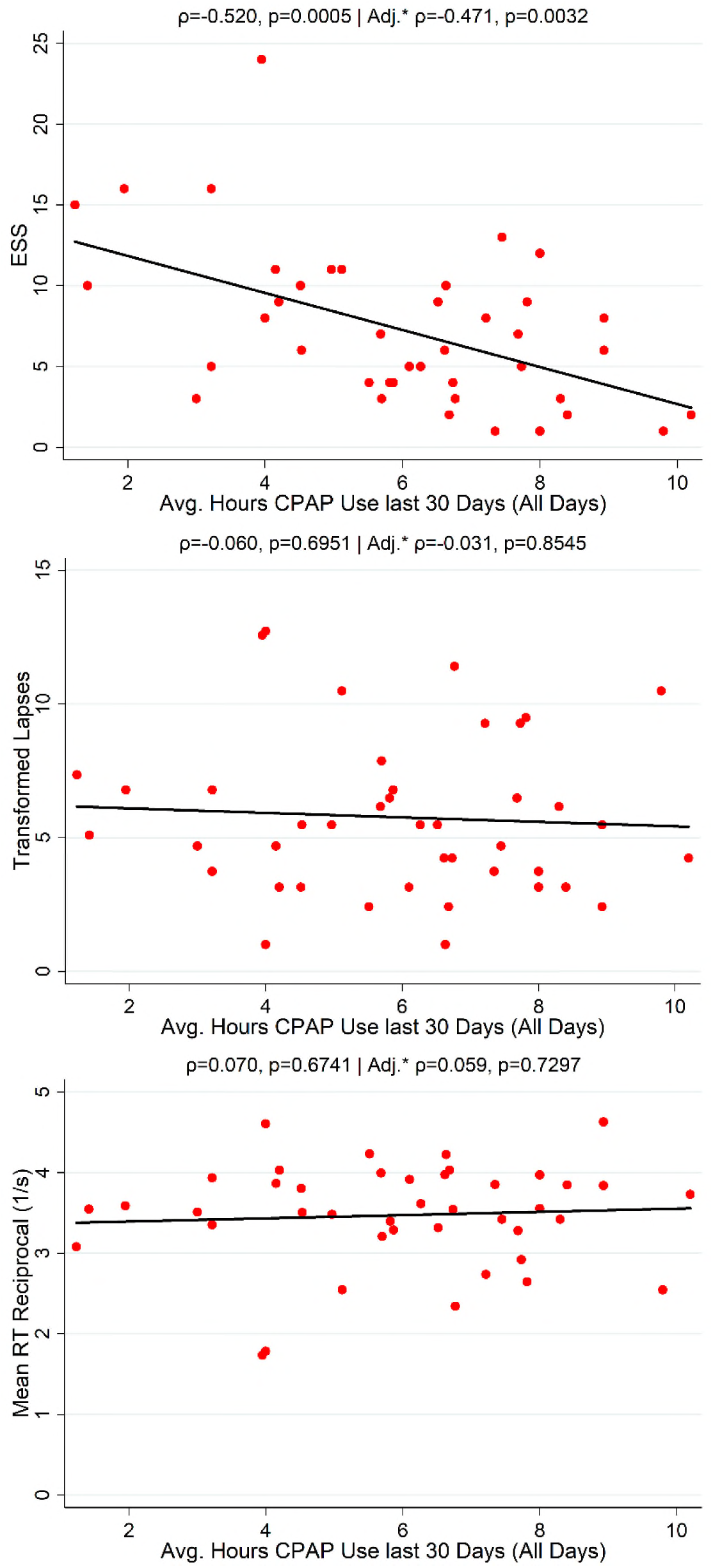
Associations between 30-day CPAP adherence and sleepiness measures among participants with OSA using CPAP. In correlation analyses, we observed a significant association between greater CPAP usage and lower ESS scores, but no association between CPAP usage and PVT measures (see also **Table 5**). *Adjusted for Age, sex and BMI.

**Table 5.** Correlations between CPAP usage and both ESS and PVT measurements.

| Sleepiness Measure | 30-day Average |  |  |  | 7-day Average |  |  |  | Previous Day |  |  |  |
| --- | --- | --- | --- | --- | --- | --- | --- | --- | --- | --- | --- | --- |
|  | *Unadjusted* |  | *Adjusted\** |  | *Unadjusted* |  | *Adjusted\** |  | *Unadjusted* |  | *Adjusted\** |  |
|  | *rho* | *p* | *rho* | *p* | *rho* | *p* | *rho* | *p* | *rho* | *p* | *rho* | *p* |
| ESS | **-0.52** | **0.001** | **-0.47** | **0.003** | **-0.47** | **0.002** | **-0.43** | **0.008** | **-0.37** | **0.016** | -0.32 | 0.0503 |
| Transformed Lapses | -0.06 | 0.695 | -0.03 | 0.855 | -0.09 | 0.551 | -0.05 | 0.789 | -0.11 | 0.501 | -0.11 | 0.538 |
| Mean RRT | 0.07 | 0.674 | 0.06 | 0.730 | 0.09 | 0.563 | 0.07 | 0.693 | 0.06 | 0.704 | 0.07 | 0.678 |
| Statistically significant associations after Hochberg correction for 3 sleepiness measures shown in **bold**. *Partial correlation adjusted for age, sex and BMI; Abbreviations: AHI = Apnea Hypopnea Index; ODI = Oxygen Desaturation Index; PVT = Psychomotor Vigilance Test; ESS = Epworth Sleepiness Scale; RRT = Reciprocal Response Time |  |  |  |  |  |  |  |  |  |  |  |  |

### Effects of CPAP Efficacy and Leak on Sleepiness

We also evaluated associations of residual AHI from the CPAP machines and device-specific leak metrics with sleepiness measures (see **Table 6**). Neither increased residual AHI nor worse leak metrics were associated with increased sleepiness on the ESS or worse performance on PVT. There was a significant, but unexpected, association between higher residual AHI and *better* performance on PVT in our data; as only 3 participants had a residual AHI over the clinically-used threshold of 10 events/hour this result is both unanticipated and likely not clinically meaningful.

**Table 6.** Correlations between CPAP efficacy and both ESS and PVT measurements.

| Sleepiness Measure | Residual AHI<sup>†</sup> |  |  |  | Time in Large Leak<sup>‡</sup> |  |  |  | 95<sup>th</sup> Percentile Leak<sup>§</sup> |  |  |  |
| --- | --- | --- | --- | --- | --- | --- | --- | --- | --- | --- | --- | --- |
|  | *Unadjusted* |  | *Adjusted\** |  | *Unadjusted* |  | *Adjusted\** |  | *Unadjusted* |  | *Adjusted\** |  |
|  | *rho* | *p* | *rho* | *p* | *rho* | *p* | *rho* | *p* | *rho* | *p* | *rho* | *p* |
| ESS | -0.11 | 0.530 | -0.20 | 0.270 | 0.49 | 0.127 | -0.09 | 0.847 | 0.13 | 0.535 | 0.13 | 0.577 |
| Transformed Lapses | -0.32 | 0.053 | **-0.51** | **0.003** | 0.04 | 0.916 | 0.10 | 0.830 | 0.12 | 0.570 | -0.12 | 0.605 |
| Mean RRT | 0.37 | 0.022 | **0.50** | **0.003** | 0.17 | 0.610 | 0.01 | 0.981 | -0.24 | 0.244 | -0.07 | 0.767 |
| Statistically significant associations after Hochberg correction for 3 sleepiness measures shown in **bold**. <sup>†</sup>Residual AHI from CPAP machine (n=38); <sup>‡</sup>Minutes in large leak available from Philips devices (n=11); <sup>§</sup>95<sup>th</sup> percentile leak (L/min) available from ResMed devices (n=25); *Partial correlation adjusted for age, sex and BMI; Abbreviations: AHI = Apnea Hypopnea Index; ODI = Oxygen Desaturation Index; PVT = Psychomotor Vigilance Test; ESS = Epworth Sleepiness Scale; RRT = Reciprocal Response Time |  |  |  |  |  |  |  |  |  |  |  |  |

## Discussion

This study reports on the relationships between subjective sleepiness based on the commonly-used Epworth Sleepiness Scale and objective attention/vigilance as measured by the Psychomotor Vigilance Test, as well as how each measure relates to OSA severity and treatment. First, we found no meaningful associations between the ESS and PVT performance, suggesting that these two measures may capture distinct aspects of the “sleepiness” reported by participants presenting to sleep clinics. Second, we observed that worse sleepiness on the ESS was associated with more severe OSA and that those with greater adherence to CPAP had less sleepiness based on the ESS. In contrast, worse PVT performance was not significantly associated with greater OSA severity or less adherence to CPAP. These data have important implications with regards to characterizing “sleepiness” or “attention” among participants presenting to sleep clinics.

The current results partially align with a prior study from Batool et al^31^ in 100 participants referred to a sleep clinic for possible sleep apnea. Daytime sleepiness was evaluated using the ESS and vigilance was measured with a portable 10-minute PVT device. The authors found that a poor PVT performance correlated with a higher ESS score, which is in line with previously published literature^30,32^, and that OSA severity measured by AHI had no correlation with PVT measures. While our study found no meaningful associations between the PVT and ESS, in contrast to this prior study, we found that OSA severity measured by AHI was correlated with ESS scores, but not with PVT performance^38,39^. Possible reasons for the differences in observed relationships between ESS and PVT in our study could be related to length of PVT used, differences in PVT devices, methods of sleep evaluation, and different patient criteria.

Ultimately, our study provides interesting new data on the relationships between subjective and objective sleepiness. As noted above, we found no meaningful associations between subjective sleepiness with the ESS and objective attention/vigilance as measured by the PVT. In other words, self-report of excessive sleepiness does not necessarily imply poor performance on PVT. This observation suggests that while these measures may be considered indicators of “sleepiness”; the two capture distinct symptoms experienced by participants; the ESS measures the tendency to fall asleep, while the PVT measures the ability to stay awake and alert^40–42^. Overall, results confirm previous studies^24,30,43–45^ suggesting that subjective and objective measures in participants with OSA should be assessed separately. These investigations^24,31,43^ have similarly found poor correlations between the PVT and subjective measures of sleepiness, leading them to conclude that objective and subjective measurements are not equivalent and should be used in conjunction for independent measures of sleep and attentional vigilance.

While our comparison of the ESS and PVT suggest these measures may capture different aspects of sleepiness, our data also suggest that the ESS tracks more closely than the PVT to OSA severity and treatment. The ESS is commonly used in sleep practices to measure sleepiness, and our data support continuing to use the ESS in evaluating participants for daytime sleepiness. We hypothesized that the PVT (since it is an objective measure) would be a better predictor of OSA severity and treatment adherence than the ESS, but our data did not support this. Contrary to our initial hypotheses, ESS scores were more strongly correlated with OSA severity than measures from PVT. Participants with greater subjective sleepiness generally had more severe OSA, but there was no meaningful relationship between OSA severity and attention/vigilance as measured by the PVT. Consistent with these observations, within a subset of participants with OSA in whom data were available, we found that more CPAP usage was associated with less sleepiness based on the ESS, but not better performance on the PVT^46^. Additional analyses showed that neither higher residual AHI nor greater leak (based on device-specific metrics) were associated with lower ESS or worse PVT performance. Therefore, making clinical recommendations based on a high residual AHI or a large mask leak at least in terms of EDS may not be clinically warranted.

Overall, these results do not strongly support a recommendation for routine use of the 3-minute PVT in participants being evaluated clinically in a sleep center. One important consideration is whether the 3-minute PVT is sufficient to capture sleepiness reported by participants in a clinical setting. It is possible that the 5 or 10-minute versions of the PVT^47,48^ may have provided more accurate measures of sustained attention and sleepiness than the 3-minute version used in our study. Evidence on the relative sensitivity of the 3-minute versus 10-minute PVT is mixed—some studies suggest that the shorter version is equally sensitive^49^, while others have found the 3-minute PVT did not produce results comparable to the 5 or 10-minute PVT in elite athletes, arguing that the two are not interchangeable^50^. Additionally, in individuals with mild OSA, the 3-minute PVT may lack the sensitivity needed to detect subtle deficits in vigilance or sleepiness. Future research could consider comparing PVT’s of longer duration to assess whether consistent results are obtained.

Another consideration is that our study examined sleepiness in a clinical population at only one point in time. Performing multiple PVT assessments over a short period (e.g., several days or even multiple times in the same day) may better characterize a patient’s attention/vigilance. Moreover, longer-term studies quantifying sleepiness using the ESS and/or PVT at multiple time points would likely better characterize the response to treatments of various sleep disorders. The lack of learning effect with the PVT^51^ likely provides a significant advantage over potential bias in self-reported measures of sleepiness with longitudinal evaluations. The technology to remotely assess PVT exists^52,53^ and is rapidly developing, although more work is needed to establish the reliability of these applications.

Having an objective longitudinal biomarker for sleepiness could better inform treatment decisions, particularly in participants in whom the self-reported symptoms are less reliable (e.g., commercial drivers^54–56^ or participants with unrecognized symptoms^19^). Repeatedly measuring such a biomarker over time could also change how we determine treatment benefits in participants treated for various sleep disorders. For instance, treatment for participants with narcolepsy is primarily based on costs and side effects, not efficacy, since there are no head-to-head trials of the commonly-used narcolepsy medications. Demonstrating objective improvements in sleepiness may facilitate an outcomes-based approach to support specific treatment options in narcolepsy and other sleep disorders. For example, does CBT-I or hypnotics in participants with insomnia or treating periodic limb movements or restless leg syndrome (RLS) improve the PVT or ESS overtime? Future studies in larger samples with these specific sleep disorders should evaluate these questions.

This study has several limitations. Most participants had never previously completed a PVT. While the task is not known to be affected by practice effects, performance may still vary due to initial unfamiliarity. A 30-second practice trial was offered before the actual test which most participants performed. Additionally, the majority of participants were being evaluated or treated for OSA, but the sample also included participants with insomnia, parasomnias, circadian rhythm disorders, and hypersomnia (narcolepsy and idiopathic hypersomnia). Future studies should include a larger population of participants with a variety of clinical sleep disorders followed longitudinally to better capture the association between subjective and objective sleepiness, including within different patient subgroups or for different conditions. While the study provides new information on the relationships between subjective and objective sleepiness within participants from an academic sleep practice, not all participants seen in the timeframe were approached or completed the PVT. This could reduce generalizability, depending on how the characteristics of included participants compare to those who did not participate. Relatedly, the sample was enriched for participants with diagnosed or suspected OSA, and results may be less generalizable to other sleep disorders. We had a relatively small sample size for certain analyses; at an α of 0.05, our sample had 80% power to detect correlations of >0.21 when comparing ESS and PVT among all participants and >0.42 when associating CPAP adherence with sleepiness measures. Thus, non-significant results for smaller correlations should be interpreted with some caution and, more generally, all results interpreted with respect to the clinical significance of associations (e.g., rho of 0.1, 0.3 and 0.5 representing small, medium and large effects^35^). Ultimately, future studies with larger sample sizes would be beneficial to both validate associations found here and to understand how sample heterogeneity influences results.

There are also important strengths that should be emphasized. The present study provides insights into the relationships between subjective and objective sleepiness among a representative (and heterogeneous) sample of participants seen at an academic clinical sleep center, enhancing generalizability. Other strengths include performing the PVT and ESS at the same visit in a diverse clinical sample, including participants with different sleep disorders (e.g., not only participants with sleep apnea), and the availability of objective CPAP adherence, efficacy (residual AHI) and mask leak data to explore associations between treatment and sleepiness in participants with OSA.

## Conclusions

In conclusion, in participants who were evaluated in an academic clinical sleep center, our data showed that the ESS tracks more closely than the PVT to OSA severity and treatment. Participants with more severe OSA demonstrated worse subjective sleepiness on the ESS, but not worse attention/vigilance based on the PVT. Similarly, amount of CPAP usage was associated with better subjective sleepiness, but not associated with better performance on PVT. In addition, there were no meaningful associations between subjective sleepiness as measured by the ESS and objective sleepiness/attention as measured by the PVT. These data suggest that the PVT and ESS may be measuring two distinct aspects of sleepiness/function experienced by participants.

## Data Availability

All data produced in the present work are contained in the manuscript.

## Abbreviations

AHI: Apnea-Hypopnea Index
BMI: Body Mass Index
CPAP: Continuous Positive Airway Pressure
EDS: Excessive Daytime Sleepiness
EHR: Electronic Health Record
ESS: Epworth Sleepiness Scale
HSAT: Home Sleep Apnea Test
OSA: Obstructive Sleep Apnea
PSG: Polysomnography
PVT: Psychomotor Vigilance Test
RRT: Reciprocal Response Time
SD: Standard Deviation

## Acknowledgments

This work was supported by the National Institute of Health grant P01 HL160471 (Developing a P4 Medicine Approach to Obstructive Sleep Apnea)

## Notes

### Competing Interest Statement

The authors have declared no competing interest.

### Funding Statement

This study did not receive any funding.

### Author Declarations

This project was reviewed and determined to qualify as Quality Improvement by the University of Pennsylvania Institutional Review Board. An IRB number was not assigned as a result.

## References

1. Lal C, Weaver TE, Bae CJ, Strohl KP. Excessive daytime sleepiness in obstructive sleep apnea. Mechanisms and clinical management. Ann Am Thorac Soc 2021;18(5):757–768. doi:10.1513/AnnalsATS.202006-696FR.

2. Johns MW. A new method for measuring daytime sleepiness: the Epworth sleepiness scale. Sleep. 1991;14(6):540–545. doi:10.1093/sleep/14.6.540.

3. Boyes J, Drakatos P, Jarrold I, Smith J, Steier J. The use of an online Epworth sleepiness scale to assess excessive daytime sleepiness. Sleep Breath. 2017;21(2):333–340. doi:10.1007/s11325-016-1417-x.

4. Seneviratne U, Puvanendran K. Excessive daytime sleepiness in obstructive sleep apnea: prevalence, severity, and predictors. Sleep Med. 2004;5(4):339–343. doi:10.1016/j.sleep.2004.01.021.

5. Feng J, He Q-y, Zhang X-l, Chen B-y, Sleep Breath Disorder Group SoRM. Epworth sleepiness scale may be an indicator for blood pressure profile and prevalence of coronary artery disease and cerebrovascular disease in patients with obstructive sleep apnea. Sleep Breath. 2012;16(1):31–40. doi:10.1007/s11325-011-0481-5.

6. Ye L, Pien GW, Ratcliffe SJ, et al. The different clinical faces of obstructive sleep apnoea: a cluster analysis. Eur Respir J. 2014;44(6):1600–7. doi:10.1183/09031936.00032314.

7. Mazzotti DR, Keenan BT, Lim DC, Gottlieb DJ, Kim J, Pack AI. Symptom subtypes of obstructive sleep apnea predict incidence of cardiovascular outcomes. Am J Respir Crit Care Med. 2019;200(4):493–506. doi:10.1164/rccm.201808-1509OC.

8. Kim J, Keenan BT, Lim DC, Lee SK, Pack AI, Shin C. Symptom-based subgroups of Koreans with obstructive sleep apnea. J Clin Sleep Med. 2018;14(3):437–443. doi:10.5664/jcsm.6994.

9. González KA, Tarraf W, Wallace DM, et al. Phenotypes of obstructive sleep apnea in the Hispanic Community Health Study/Study of Latinos. Sleep. 2021;44(12)doi:10.1093/sleep/zsab181.

10. Allen AJH, Jen R, Mazzotti DR, et al. Symptom subtypes and risk of incident cardiovascular and cerebrovascular disease in a clinic-based obstructive sleep apnea cohort. J Clin Sleep Med. 2022;18(9):2093–2102. doi:10.5664/jcsm.9986.

11. Gervès-Pinquié C, Bailly S, Goupil F, et al. Positive airway pressure adherence, mortality, and cardiovascular events in patients with sleep apnea. Am J Respir Crit Care Med. 2022;206(11):1393–1404. doi:10.1164/rccm.202202-0366OC.

12. Rosenthal LD, Dolan DC. The Epworth sleepiness scale in the idenfificafion of obstructive sleep apnea. J Nerv Ment Dis. 2008;196(5):429–431. doi:10.1097/NMD.0b013e31816ff3bf.

13. Guo Q, Song W-d, Li W, et al. Weighted Epworth sleepiness scale predicted the apnea-hypopnea index better. Respir Res. 2020;21(1):147. doi:10.1186/s12931-020-01417-w.

14. Hsieh P-S, Hwang S-W, Hwang S-R, Hwang J-H. Association between various breathing indexes during sleep and the Epworth sleepiness scale score in adults. Med. 2022;101(48):e32017. doi:10.1097/md.0000000000032017.

15. Thorarinsdottir EH, Pack AI, Gislason T, et al. Polysomnographic characteristics of excessive daytime sleepiness phenotypes in obstructive sleep apnea: results from the international sleep apnea global interdisciplinary consortium. Sleep. 2024;47(4)doi:10.1093/sleep/zsae035.

16. Keenan BT, Kim J, Singh B, et al. Recognizable clinical subtypes of obstructive sleep apnea across international sleep centers: a cluster analysis. Sleep. 2018;41(3)doi:10.1093/sleep/zsx214.

17. Allen AH, Beaudin AE, Fox N, et al. Symptom subtypes and cognitive function in a clinic-based OSA cohort: a multi-centre Canadian study. Sleep Med. 2020;74:92–98. doi:10.1016/j.sleep.2020.05.001.

18. Trzepizur W, Blanchard M, Ganem T, et al. Sleep apnea-specific hypoxic burden, symptom subtypes, and risk of cardiovascular events and all-cause mortality. Am J Respir Crit Care Med. 2022;205(1):108–117. doi:10.1164/rccm.202105-1274OC.

19. Keenan BT, Ye L, Pien GW, et al. Symptom Subtypes of Obstructive Sleep Apnea 10 Years Later: Past, Present and Future. Sleep. Apr 9 2025;doi:10.1093/sleep/zsaf082

20. Mazzotti DR, Keenan BT, Thorarinsdottir EH, Gislason T, Pack AI. Is the Epworth sleepiness scale sufficient to identify the excessively sleepy subtype of OSA? Chest. 2022;161(2):557–561. doi:10.1016/j.chest.2021.10.027.

21. Dinges DF, Powell JW. Microcomputer analyses of performance on a portable, simple visual RT task during sustained operations. Behav Res Methods. 1985;17(6):652–655. doi:10.3758/BF03200977.

22. Basner M, Dinges DF. Maximizing sensitivity of the psychomotor vigilance test (PVT) to sleep loss. Sleep. 2011;34(5):581–91. doi:10.1093/sleep/34.5.581.

23. Jones CW, Basner M, Mollicone DJ, Mott CM, Dinges DF. Sleep deficiency in spaceflight is associated with degraded neurobehavioral functions and elevated stress in astronauts on six-month missions aboard the International Space Station. Sleep. 2022;45(3)doi:10.1093/sleep/zsac006.

24. Zhang C, Varvarigou V, Parks PD, et al. Psychomotor vigilance testing of professional drivers in the occupational health clinic: a potential objective screen for daytime sleepiness. J Occup Environ Med. 2012;54(3):296–302. doi:10.1097/JOM.0b013e318223d3d6.

25. Behrens T, Burek K, Pallapies D, et al. Decreased psychomotor vigilance of female shift workers after working night shifts. PLoS One. 2019;14(7):e0219087. doi:10.1371/journal.pone.0219087.

26. Surani S, Hesselbacher S, Guntupalli B, Surani S, Subramanian S. Sleep quality and vigilance differ among inpatient nurses based on the unit setting and shift worked. J Patient Saf. 2015;11(4):215–220. doi:10.1097/pts.0000000000000089.

27. Basner M, Mollicone D, Dinges DF. Validity and sensitivity of a brief psychomotor vigilance test (PVT-B) to total and partial sleep deprivation. Acta Astronaut. 2011;69(11-12):949–959. doi:10.1016/j.actaastro.2011.07.015.

28. Thomann J, Baumann CR, Landolt HP, Werth E. Psychomotor vigilance task demonstrates impaired vigilance in disorders with excessive daytime sleepiness. J Clin Sleep Med. 2014;10(9):1019–24. doi:10.5664/jcsm.4042.

29. Lim J, Dinges DF. Sleep deprivation and vigilant aftenfion. Ann N Y Acad Sci. 2008;1129(1):305–322. doi:10.1196/annals.1417.002.

30. Li Y, Vgontzas A, Kritikou I, et al. Psychomotor vigilance test and its association with daytime sleepiness and inflammation in sleep apnea: clinical implications. J Clin Sleep Med. Sep 15 2017;13(9):1049–1056. doi:10.5664/jcsm.6720.

31. Batool-Anwar S, Kales SN, Patel SR, Varvarigou V, DeYoung PN, Malhotra A. Obstructive sleep apnea and psychomotor vigilance task performance. Nat Sci Sleep. 2014;6:65–71. doi:10.2147/nss.S53721.

32. Kim H, Dinges DF, Young T. Sleep-disordered breathing and psychomotor vigilance in a community-based sample. Sleep. 2007;30(10):1309–1316. doi:10.1093/sleep/30.10.1309.

33. Schwab RJ, Badr SM, Epstein LJ, et al. An official American Thoracic Society statement: continuous positive airway pressure adherence tracking systems. The optimal monitoring strategies and outcome measures in adults. Am J Respir Crit Care Med. Sep 1 2013;188(5):613–20. doi:10.1164/rccm.201307-1282ST

34. Basner M, Rubinstein J. Fitness for duty: a 3-minute version of the Psychomotor Vigilance Test predicts fatigue-related declines in luggage-screening performance. J Occup Environ Med. 2011;53(10):1146–54. doi:10.1097/JOM.0b013e31822b8356.

35. Cohen J. Statistical power analysis for the behavioral sciences 2nd ed. ed. 1988.

36. Hochberg Y. A sharper Bonferroni procedure for multiple tests of significance. Biometrika. 1988;75(4):800–802. doi:10.1093/biomet/75.4.800.

37. Huang Y, Hsu JC. Hochberg’s step-up method: cutting corners off holm’s step-down method. Biometrika. 2007;94(4):965–975. doi:10.1093/biomet/asm067.

38. Lee IS, Bardwell WA, Ancoli-Israel S, Dimsdale JE. Number of lapses during the psychomotor vigilance task as an objective measure of fatigue. J Clin Sleep Med. 2010;6(2):163–8.

39. Pack AI, Maislin G, Staley B, et al. Impaired performance in commercial drivers: role of sleep apnea and short sleep duration. Am J Respir Crit Care Med. 2006;174(4):446–454. doi:10.1164/rccm.200408-1146OC.

40. Huang Y, Hennig S, Fietze I, Penzel T, Veauthier C. The psychomotor vigilance test compared to a divided aftenfion steering simulation in patients with moderate or severe obstructive sleep apnea. Nat Sci Sleep. 2020;12:509–524. doi:10.2147/nss.S256987.

41. Sunwoo BY, Jackson N, Maislin G, Gurubhagavatula I, George CF, Pack AI. Reliability of a single objective measure in assessing sleepiness. Sleep. 2012;35(1):149–158. doi:10.5665/sleep.1606.

42. Johns M. Rethinking the assessment of sleepiness. Sleep Med Rev. 1998;2(1):3–15. doi:10.1016/S1087-0792(98)90050-8.

43. Bhat S, Gupta D, Akel O, et al. The relationships between improvements in daytime sleepiness, fatigue and depression and psychomotor vigilance task testing with CPAP use in patients with obstructive sleep apnea. Sleep Med. 2018;49:81–89. doi:10.1016/j.sleep.2018.06.012.

44. Harel BT, Plazzi G, Pizza F, et al. Dissociation of symptoms of excessive daytime sleepiness and aftenfional impairment in narcolepsy type 1. presented at: Sleep Europe 2024; Seville, Spain.

45. Franzen PL, Siegle GJ, Buysse DJ. Relationships between affect, vigilance, and sleepiness following sleep deprivation. J Sleep Res. 2008;17(1):34–41. doi:10.1111/j.1365-2869.2008.00635.x

46. Weaver TE, Maislin G, Dinges DF, et al. Relationship between hours of CPAP use and achieving normal levels of sleepiness and daily functioning. Sleep. Jun 2007;30(6):711–9. doi:10.1093/sleep/30.6.711

47. Roach GD, Dawson D, Lamond N. Can a shorter psychomotor vigilance task be used as a reasonable substitute for the ten-minute psychomotor vigilance task? Chronobiol Int. 2006;23(6):1379–87. doi:10.1080/07420520601067931

48. Thompson BJ, Shugart C, Dennison K, Louder TJ. Test-retest reliability of the 5-minute psychomotor vigilance task in working-aged females. J Neurosci Methods. Jan 1 2022;365:109379. doi:10.1016/j.jneumeth.2021.109379

49. Benderoth S, Hörmann HJ, Schießl C, Elmenhorst EM. Reliability and validity of a 3-min psychomotor vigilance task in assessing sensitivity to sleep loss and alcohol: fitness for duty in aviation and transportation. Sleep. 2021;44(11)doi:10.1093/sleep/zsab151

50. Jones MJ, C. DI, Kevin M, et al. The psychomotor vigilance test: a comparison of different test durations in elite athletes. J Sports Sci. 2018/09/17 2018;36(18):2033–2037. doi:10.1080/02640414.2018.1433443

51. Basner M, Hermosillo E, Nasrini J, et al. Repeated administration effects on psychomotor vigilance test performance. Sleep. 2017;41(1)doi:10.1093/sleep/zsx187.

52. Arsintescu L, Mulligan JB, Flynn-Evans EE. Evaluation of a psychomotor vigilance task for touch screen devices. Hum Factors Health. 2017;59(4):661–670. doi:10.1177/0018720816688394.

53. Arsintescu L, Kato KH, Cravalho PF, Feick NH, Stone LS, Flynn-Evans EE. Validation of a touchscreen psychomotor vigilance task. Accid Anal Prev. 2019;126:173–176. doi:10.1016/j.aap.2017.11.041.

54. Kneeland E, Ali N, Maislin DG, et al. Achieving adherence to positive airway pressure in commercial drivers using an employer-mandated remote management programme. ERJ Open Res. 2024;10(6):00132–2024. doi:10.1183/23120541.00132-2024

55. Lyons MM, Kraemer JF, Dhingra R, et al. Screening for Obstructive Sleep Apnea in Commercial Drivers Using EKG-Derived Respiratory Power Index. J Clin Sleep Med. Jan 15 2019;15(1):23–32. doi:10.5664/jcsm.7562

56. Howard ME, Desai AV, Grunstein RR, et al. Sleepiness, sleep-disordered breathing, and accident risk factors in commercial vehicle drivers. Am J Respir Crit Care Med. 2004;170(9):1014–21. doi:10.1164/rccm.200312-1782OC

